# Fully Vaccinated and Boosted Patients Requiring Hospitalization for COVID-19: an Observational Cohort Analysis

**DOI:** 10.1101/2022.01.05.22268626

**Authors:** Nicholas Mielke, Steven Johnson, Amit Bahl

## Abstract

**Objective:** Real-world data on the effectiveness of boosters against COVID-19, especially as new variants continue to emerge, is limited. It is our objective to assess demographic, clinical, and outcome variables of patients requiring hospitalization for severe SARS-CoV-2 infection comparing fully vaccinated and boosted (FV&B) and unvaccinated (UV) patients.

**Methods:** This multicenter observational cohort analysis compared demographic, clinical, and outcome variables in FV&B and UV adults hospitalized for COVID-19. A sub-analysis of FV&B patients requiring intensive care (ICU) care versus non-ICU care was performed to describe and analyze common symptom presentations, initial vital signs, initial laboratory workup, and pertinent medication use in these two groups.

**Results:** Between August 12^th^, 2021 and December 6^th^, 2021, 4,571 patient encounters had a primary diagnosis of COVID-19 and required inpatient treatment at an acute-care hospital system in Southeastern Michigan. Of the 4,571 encounters requiring hospitalization, 65(1.4%) were FV&B and 2,935(64%) were UV. FV&B individuals were older (74 [67, 81] vs 58 [45, 70]; p <0.001) with a higher proportion of immunocompromised individuals (32.3% vs 10.4%; p<0.001). Despite a significantly higher baseline risk of in-hospital mortality in the FV&B group compared to the UV (Elixhauser 16 vs 8 (p <0.001)), there was a trend toward lower in-hospital mortality (7.7% vs 12.1%; p=0.38) among FV&B patients. Other severe outcomes followed this same trend, with 7.7% of FV&B vs 11.1% UV patients needing mechanical ventilation and 4.6% vs 10.6% of patients needing vasopressors in each group, respectively (p=0.5 and 0.17).

**Conclusions:** Fully vaccinated and boosted individuals requiring hospital-level care for breakthrough COVID-19 tended to have less severe outcomes despite appearing to be higher risk at baseline when compared to unvaccinated individuals during the same time period. Specifically, there was a trend that FV&B group had lower rates of mechanical ventilation, use of vasopressors, and in-hospital mortality. As COVID-19 continues to spread, larger expansive trials are needed to further identify risk factors for severe outcomes among the FV&B population.

## INTRODUCTION

The Coronavirus Disease (COVID-19) pandemic landscape has changed dramatically since it began.^1^ In just one year, scientists, industry, and the governments were able to develop and distribute safe and effective vaccinations that offer protection against the SARS-CoV-2 virus.^2–4^ However, despite these advances, the death toll has been substantial, with nearly 1 in 100 Americans over the age of 65 succumbing to this deadly virus.^5^ As SARS-CoV-2 infection continues to spread around the globe, one critical challenge is the continued viral mutation leading to new and potentially dangerous viral variants that may decrease the efficacy of current vaccines. Fortunately, studies on the B.1.351, B.1.1.7 (alpha), and B.1.617.2 (delta) variants revealed no major decrease in effectiveness with the Pfizer-BioNTech vaccine.^6,7^ However, as new variants continue to emerge, breakthrough SARS-CoV-2 infections in vaccinated individuals remain a significant concern. Smaller studies on fully vaccinated healthcare workers who subsequently developed SARS-CoV-2 infections demonstrated rates of breakthrough infection between 0.05% - 11.6%.^8–11^.

Early data from Israel and the United Kingdom also demonstrated that the effectiveness of the vaccine for preventing breakthrough infection and severe disease waned as time from vaccination increased.^12,13^ In order to combat this potential for waning immunity, the United States Food and Drug Administration authorized booster vaccinations in all individuals who completed the primary vaccination series at least six months after receiving the Moderna or Pfizer-BioNTech SARS-CoV-2 vaccine or at least two months after being vaccinated with the Janssen SARS-CoV-2 vaccine.^14,15^ To date, there is minimal data regarding the effectiveness of a booster dose in preventing breakthrough COVID-19 and progression to severe disease, especially in areas with a high concentration of viral variants. It is our objective to assess shared characteristics of patients requiring hospitalization for severe SARS-CoV-2 infection comparing fully vaccinated and boosted (FV&B) and unvaccinated (UV) patients.

## MATERIALS AND METHODS

### Study Design, Setting, and Participants

We conducted a multicenter observational cohort analysis evaluating adult patients requiring hospital admission for COVID-19. We compared demographic and health history data between patients who had completed a SARS-CoV-2 primary vaccination series and also received a booster dose (FV&B) with all unvaccinated (UV) patients who required hospitalization for COVID-19 during the same time period. Further, we analyzed clinical variables within the FV&B population to identify which variables at presentation to the Emergency Department (ED) correlated with need for intensive care unit (ICU) care. Patients were identified through analysis of electronic health records (EHR; Epic Systems, Verona, WI, USA). This study was conducted at Beaumont Health, an eight-hospital acute care hospital system in southeastern Michigan.

Consecutive adult patients greater than or equal to 18 years of age presenting to the ED and requiring hospital admission with a discharge diagnosis indicating active COVID-19 disease were eligible for inclusion. Patients who had laboratory-confirmed SARS-CoV-2 infection during a previous hospitalization and pediatric patients were excluded. The study was approved by the home organization’s Institutional Review Board. Written informed consent requirement was waived.

### Study Definitions

Patients were categorized as either UV or FV&B. UV individuals had no record of immunization against SARS-CoV-2 available in our EHR or within the Michigan Care Improvement Registry (MICR), which tracks all vaccinations from any source across the state of Michigan.^16^ Per the current Centers for Disease Control and Prevention (CDC) guidelines, FV&B individuals had received either three doses of an mRNA vaccine (Pfizer, Moderna), one dose of a viral vector vaccine (Janssen) and one dose of the mRNA, one dose of a viral vector vaccine and two doses of the mRNA, or two doses of a viral vector vaccine.^17^ All individuals who received the correct number and combination of doses were included in the analysis irrespective of the timeline of dose administration. In-hospital mortality included death during the hospital stay as well as death within ten days of being discharged to hospice.

### Data sources/measurement

All data were extracted through the use of the EHR. This data included demographic, clinical, laboratory, and outcomes variables. Additionally, data included comorbidities, number of ED visits in the past six months, and body mass index (BMI) was extracted from the EHR after our cohort was identified. Comorbidities were assessed using the Agency for Healthcare Research and Quality (AHRQ) Elixhauser Comorbidity Index.^18^ Specific to the FV&B group, we extracted initial ED vital signs and laboratory values. A manual chart review was performed to identify the time from symptom onset to ED presentation. For both cohorts, we evaluated hospital treatment and outcomes data. These variables included: need for supplemental oxygen stratified by level of support including mechanical ventilation, highest level of care, need for renal replacement therapy, vasopressor use, extracorporeal membrane oxygenation (ECMO) use, hospital length of stay, and hospital disposition.

Data from the MCIR included dates of vaccine administration as well as vaccine type and brand.

Immunocompromised individuals were identified using the AHRQ list of ICD-10 diagnoses and procedure codes that they have listed as defining an immunocompromised state (IMMUNID).^19^ All available EHR data were used to screen for these conditions and procedures and codes that were deemed active during the current hospitalization were included.

Anticoagulants in this study included oral Apixaban, subcutaneous Enoxaparin, Heparin (subcutaneous or intravenous), oral Rivaroxaban, and oral Warfarin. Corticosteroids included Dexamethasone (oral or intravenous), oral Prednisone, and intravenous Hydrocortisone succinate.

### Outcome Measures

The primary outcome of this study was the comparison of demographic and health history data between FV&B individuals who were hospitalized for breakthrough COVID-19 to UV individuals who were hospitalized for COVID-19 during the same time period. Secondary outcomes included the analysis of clinical variables within the FV&B population to identify if any variables at ED presentation that were different amongst patients who required ICU level of care versus those who did not.

### Statistical Analysis

All data were summarized through descriptive statistics. Numerical results are reported as means and standard deviation or medians and interquartile ranges. Categorical variables are reported as counts and percentages. The Chi-squared or Fisher’s exact test (categorical variables) and the Kruskal-Wallis exact test (continuous variables) were used to compare differences among FV&B and UB as well as FV&B ICU patients and FV&B non-ICU patients. All tests performed in this analysis were two-sided tests, with p < 0.05 or a confidence interval of 95% indicating statistical significance. Analysis was performed using R software, version 4.1.2 (R Foundation for Statistical Computing) and Excel (Microsoft).

## RESULTS

Between August 12^th^, 2021 and December 6^th^, 2021, there were 10,236 ED encounters with a primary diagnosis of COVID-19 (ICD 10: U07.1). After excluding 5,665 encounters that were discharged from the ED, 1242 fully vaccinated but not boosted encounters, and 329 partially vaccinated encounters, a total of 3000 encounters remained in the final analysis. Of these cases, 65 (2.2%) were FV&B and 2935 (97.8%) were UV. **(Figure 1)**

**Figure 1.**
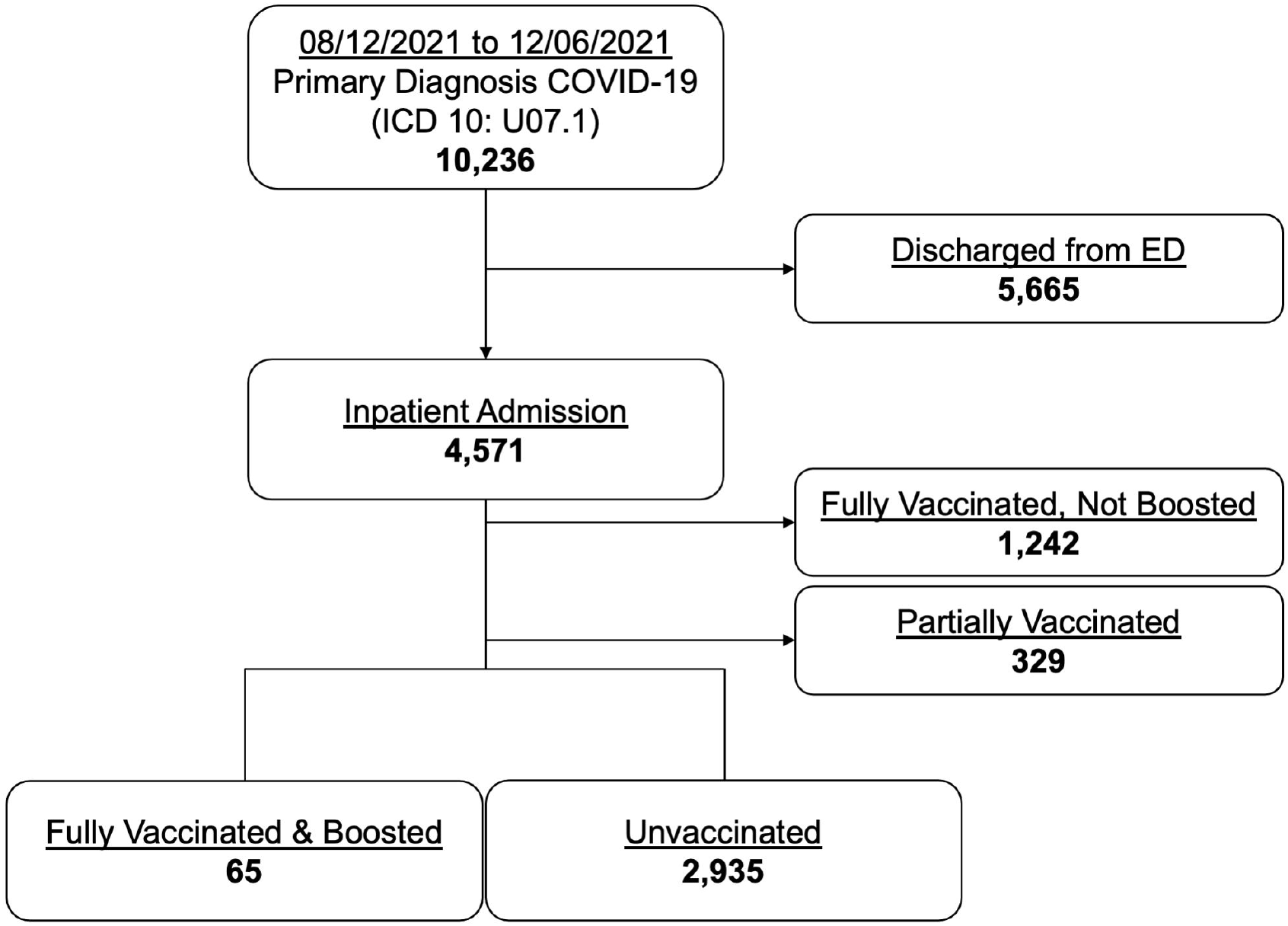
Enrollment profile of COVID-19 emergency and inpatient encounters.

The median age of FV&B was 74 (67,81) compared to 58 (45,70) for UV (p<0.001). Majority were female in both the FB&V and UV groups with 56.9% and 52.5%, respectively. FV&B had a lower BMI 29.7 (24.2, 33.8) compared to UV 30.2 (25.9, 35.9) (p=0.08). The proportion of immunocompromised and pre-existing end-stage renal disease (ESRD) in the FV&B cohort were 32.3% and 18.5%, respectively, compared to 10.4% and 1.8% in the UV cohort, respectively (p<0.001 for both). The Elixhauser weighted score was 16 (6,26) for FV&B and 8 (0,16) for UV groups (p<0.001). 7.7% of FV&B individuals required mechanical ventilation compared to 11.1% of UV. In-hospital mortality was 7.7% in the FV&B group compared to 12.1% in the UV group (p=0.38) **(Table 1)** In a sub-analysis comparing FV&B (n=46) and UV (n=1046) patients > 65 years old, outcome differences between groups were more apparent. Specifically, in-hospital mortality was 6.5% in FV&B compared to 20.2% in UV group (p=0.08). **(Table 2)**

**Table 1.** Demographics, pre-existing comorbidities, in-hospital therapies, and outcomes among FV&B and UV individuals.

| Variables<sup>‡</sup> | FV&B | UV | *p* value |
| --- | --- | --- | --- |
| n | 65 | 2935 |  |
| Demographics |  |  |  |
| Age, years |  |  |  |
| Mean (SD) | 72.06 (12.09) | 57.63 (17.07) | <0.001<sup>¶</sup> |
| Median (IQRs) | 74.00 (67.00, 81.00) | 58.00 (45.00, 70.00) |  |
| Gender |  |  |  |
| Female | 37/65 (56.92%) | 1,541/2,935 (52.50%) | 0.562<sup>‡</sup> |
| Male | 28/65 (43.08%) | 1,394/2,935 (47.50%) |  |
| Race |  |  |  |
| American Indian or Alaska Native | 0/65 (0.00%) | 6/2,935 (0.20%) | 0.437<sup>‡</sup> |
| Asian | 0/65 (0.00%) | 23/2,935 (0.78%) |  |
| Black or African American | 9/65 (13.85%) | 704/2,935 (23.99%) |  |
| Native Hawaiian/Pacific Islander | 0/65 (0.00%) | 2/2,935 (0.07%) |  |
| White or Caucasian | 53/65 (81.54%) | 2,029/2,935 (69.13%) |  |
| Other | 3/65 (4.62%) | 161/2,935 (5.49%) |  |
| Not available | 0/65 (0.00%) | 10/2,935 (0.34%) |  |
| BMI, kg/m<sup>2</sup> (n=65/2877) |  |  |  |
| Mean | 29.49 (7.04) | 31.71 (8.55) | 0.084<sup>¶</sup> |
| Median | 29.68 (24.20, 33.79) | 30.23 (25.94, 35.93) |  |
| Pre-existing comorbidities |  |  |  |
| ESRD | 12/65 (18.46%) | 54/2,935 (1.84%) | <0.001<sup>‡</sup> |
| Immunocompromised | 21/65 (32.31%) | 306/2,935 (10.43%) | <0.001<sup>‡</sup> |
| Elixhauser weighted score (n=65/2934) |  |  |  |
| Mean | 16.02 (12.58) | 9.00 (11.26) | <0.001<sup>‡</sup> |
| Median | 16.00 (6.00, 26.00) | 8.00 (0.00, 16.00) |  |
| In-hospital therapies |  |  |  |
| O<sub>2</sub> therapy | 48/65 (73.85%) | 2,331/2,935 (79.42%) | 0.346<sup>‡</sup> |
| Nasal cannula/non-rebreather | 30/65 (46.15%) | 1,156/2,935 (39.39%) | 0.329<sup>‡</sup> |
| High flow O<sub>2</sub> | 6/65 (9.23%) | 612/2,935 (20.85%) | 0.033<sup>‡</sup> |
| Non-invasive ventilation | 7/65 (10.77%) | 237/2,935 (8.07%) | 0.578<sup>‡</sup> |
| Mechanical ventilation | 5/65 (7.69%) | 326/2,935 (11.11%) | 0.503<sup>‡</sup> |
| Vasopressor | 3/65 (4.62%) | 312/2,935 (10.63%) | 0.174<sup>‡</sup> |
| ICU-level care | 11/65 (16.92%) | 493/2,935 (16.80%) | 1.000<sup>‡</sup> |
| Length of stay, hours (n=64/2858) |  |  |  |
| Mean | 192.03 (205.92) | 186.63 (169.35) | 0.870<sup>¶</sup> |
| Median | 133.50 (79.00, 256.00) | 138.00 (76.00, 240.00) |  |
| Outcomes |  |  |  |
| Death | 5/65 (7.69%) | 354/2,935 (12.06%) | 0.379<sup>‡</sup> |
| Discharge disposition location |  |  |  |
| Home | 49/60 (81.67%) | 2,277/2,581 (88.22%) | 0.058<sup>‡</sup> |
| Rehab | 3/60 (5.00%) | 27/2,581 (1.05%) |  |
| SNF | 8/60 (13.33%) | 188/2,581 (7.28%) |  |
| Transferred | 0/60 (0.00%) | 15/2,581 (0.58%) |  |
| Hospice | 0/60 (0.00%) | 11/2,581 (0.43%) |  |
| Unknown | 0/60 (0.00%) | 63/2,581 (2.44%) |  |

Abbreviations: FV&B=fully vaccinated and boosted; UV=unvaccinated; BMI=body mass index; ESRD=end-stage renal disease; ICU=intensive care unit; Rehab=rehabilitation; SNF=skilled nursing facility
<sup>‡</sup>For continuous variables, medians (interquartile ranges, IQRs) and means (standard deviation, SD) were presented. For categorical variables, frequencies (percentage) were presented.
<sup>¶</sup>Kruskal-Wallis test
<sup>‡</sup>Chi-squared or Fisher's exact test

**Table 2.**
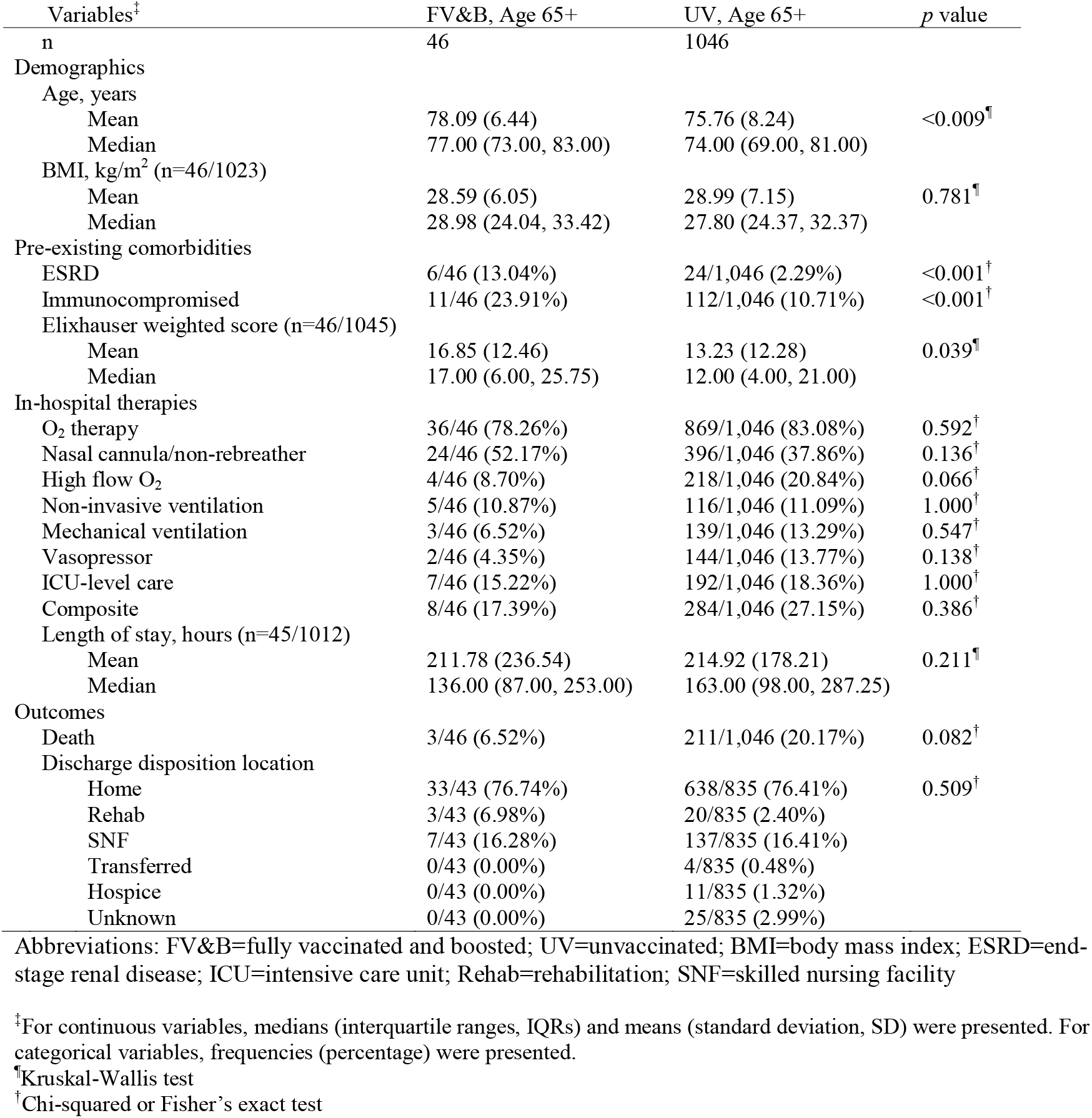
Demographics, pre-existing comorbidities, in-hospital therapies, and outcomes among individuals ages 65 and older in the FV&B and UV cohorts

| Variables<sup>‡</sup><br>n | FV&B, Age 65+<br>46 | UV, Age 65+<br>1046 | p value |
| --- | --- | --- | --- |
| Demographics |  |  |  |
| Age, years |  |  |  |
| Mean | 78.09 (6.44) | 75.76 (8.24) | <0.009<sup>¶</sup> |
| Median | 77.00 (73.00, 83.00) | 74.00 (69.00, 81.00) |  |
| BMI, kg/m<sup>2</sup> (n=46/1023) |  |  |  |
| Mean | 28.59 (6.05) | 28.99 (7.15) | 0.781<sup>¶</sup> |
| Median | 28.98 (24.04, 33.42) | 27.80 (24.37, 32.37) |  |
| Pre-existing comorbidities |  |  |  |
| ESRD | 6/46 (13.04%) | 24/1,046 (2.29%) | <0.001<sup>†</sup> |
| Immunocompromised | 11/46 (23.91%) | 112/1,046 (10.71%) | <0.001<sup>†</sup> |
| Elixhauser weighted score (n=46/1045) |  |  |  |
| Mean | 16.85 (12.46) | 13.23 (12.28) | 0.039<sup>¶</sup> |
| Median | 17.00 (6.00, 25.75) | 12.00 (4.00, 21.00) |  |
| In-hospital therapies |  |  |  |
| O<sub>2</sub> therapy | 36/46 (78.26%) | 869/1,046 (83.08%) | 0.592<sup>†</sup> |
| Nasal cannula/non-rebreather | 24/46 (52.17%) | 396/1,046 (37.86%) | 0.136<sup>†</sup> |
| High flow O<sub>2</sub> | 4/46 (8.70%) | 218/1,046 (20.84%) | 0.066<sup>†</sup> |
| Non-invasive ventilation | 5/46 (10.87%) | 116/1,046 (11.09%) | 1.000<sup>†</sup> |
| Mechanical ventilation | 3/46 (6.52%) | 139/1,046 (13.29%) | 0.547<sup>†</sup> |
| Vasopressor | 2/46 (4.35%) | 144/1,046 (13.77%) | 0.138<sup>†</sup> |
| ICU-level care | 7/46 (15.22%) | 192/1,046 (18.36%) | 1.000<sup>†</sup> |
| Composite | 8/46 (17.39%) | 284/1,046 (27.15%) | 0.386<sup>†</sup> |
| Length of stay, hours (n=45/1012) |  |  |  |
| Mean | 211.78 (236.54) | 214.92 (178.21) | 0.211<sup>¶</sup> |
| Median | 136.00 (87.00, 253.00) | 163.00 (98.00, 287.25) |  |
| Outcomes |  |  |  |
| Death | 3/46 (6.52%) | 211/1,046 (20.17%) | 0.082<sup>†</sup> |
| Discharge disposition location |  |  |  |
| Home | 33/43 (76.74%) | 638/835 (76.41%) | 0.509<sup>†</sup> |
| Rehab | 3/43 (6.98%) | 20/835 (2.40%) |  |
| SNF | 7/43 (16.28%) | 137/835 (16.41%) |  |
| Transferred | 0/43 (0.00%) | 4/835 (0.48%) |  |
| Hospice | 0/43 (0.00%) | 11/835 (1.32%) |  |
| Unknown | 0/43 (0.00%) | 25/835 (2.99%) |  |

Abbreviations: FV&B=fully vaccinated and boosted; UV=unvaccinated; BMI=body mass index; ESRD=end-stage renal disease; ICU=intensive care unit; Rehab=rehabilitation; SNF=skilled nursing facility
<sup>‡</sup>For continuous variables, medians (interquartile ranges, IQRs) and means (standard deviation, SD) were presented. For categorical variables, frequencies (percentage) were presented.
<sup>¶</sup>Kruskal-Wallis test
<sup>†</sup>Chi-squared or Fisher's exact test

The majority, 59 (90.8%), of the FV&B had documentation of upper respiratory symptoms (cough with or without sputum production, shortness of breath, and/or sore throat) or systemic symptoms (fever or fatigue). 55 (84.6%) presented with respiratory symptoms, 43 (66.2%) presented with systemic symptoms, and 35 (53.8%) presented with fatigue. Regarding vaccination type, 37 (56.9%) were vaccinated with three consecutive Pfizer immunizations, 21 (32.3%) received three doses of the Moderna vaccine, and 5 (7.7%) received 2 Janssen vaccines. 27 of 65 (42%) patients experienced COVID-19 symptoms <12 days after receiving their booster vaccine. Of the 11 patients who required ICU care, 5 (45%), had symptom onset <12 days after receiving their booster dose. **(Table 3)**

**Table 3.**
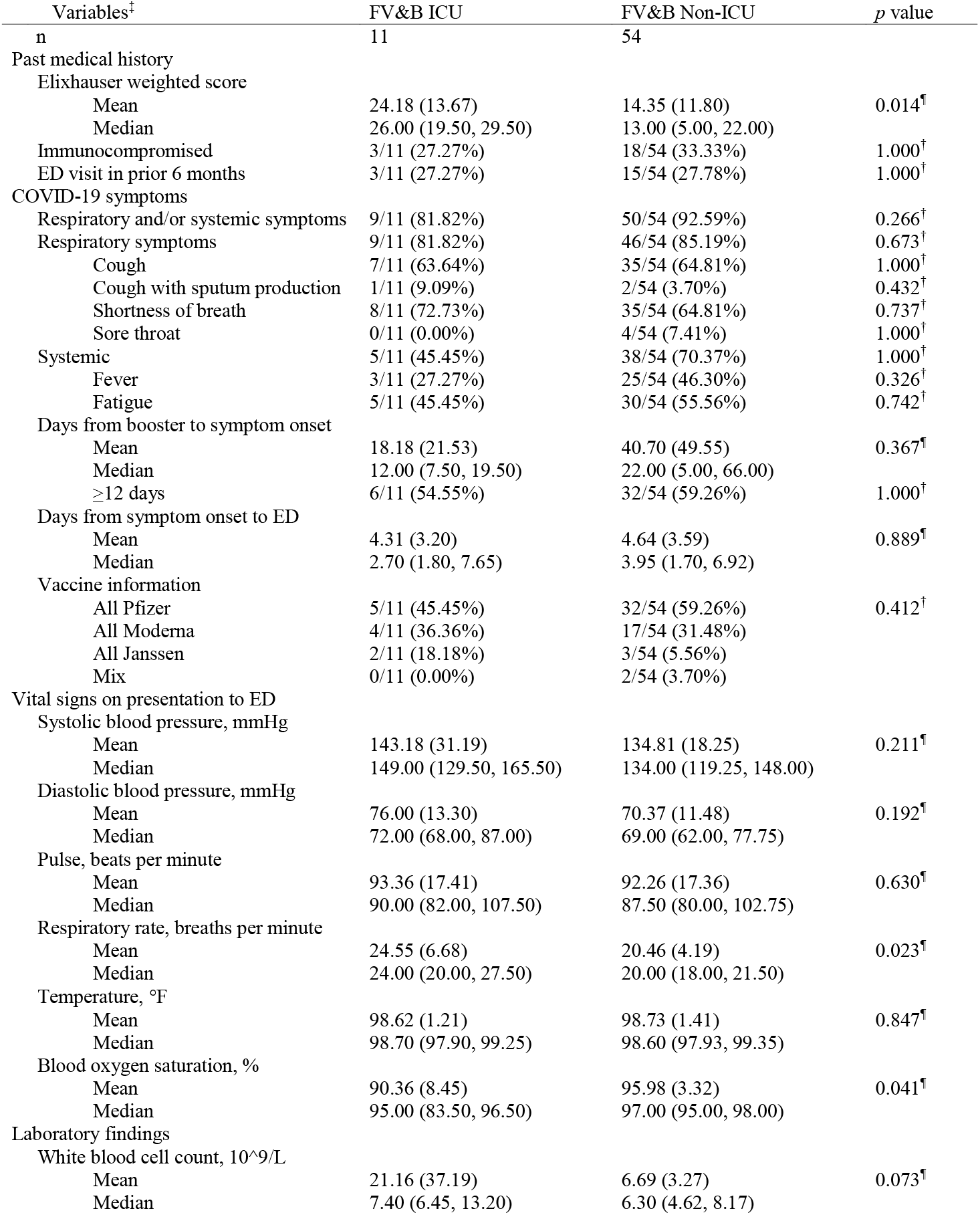
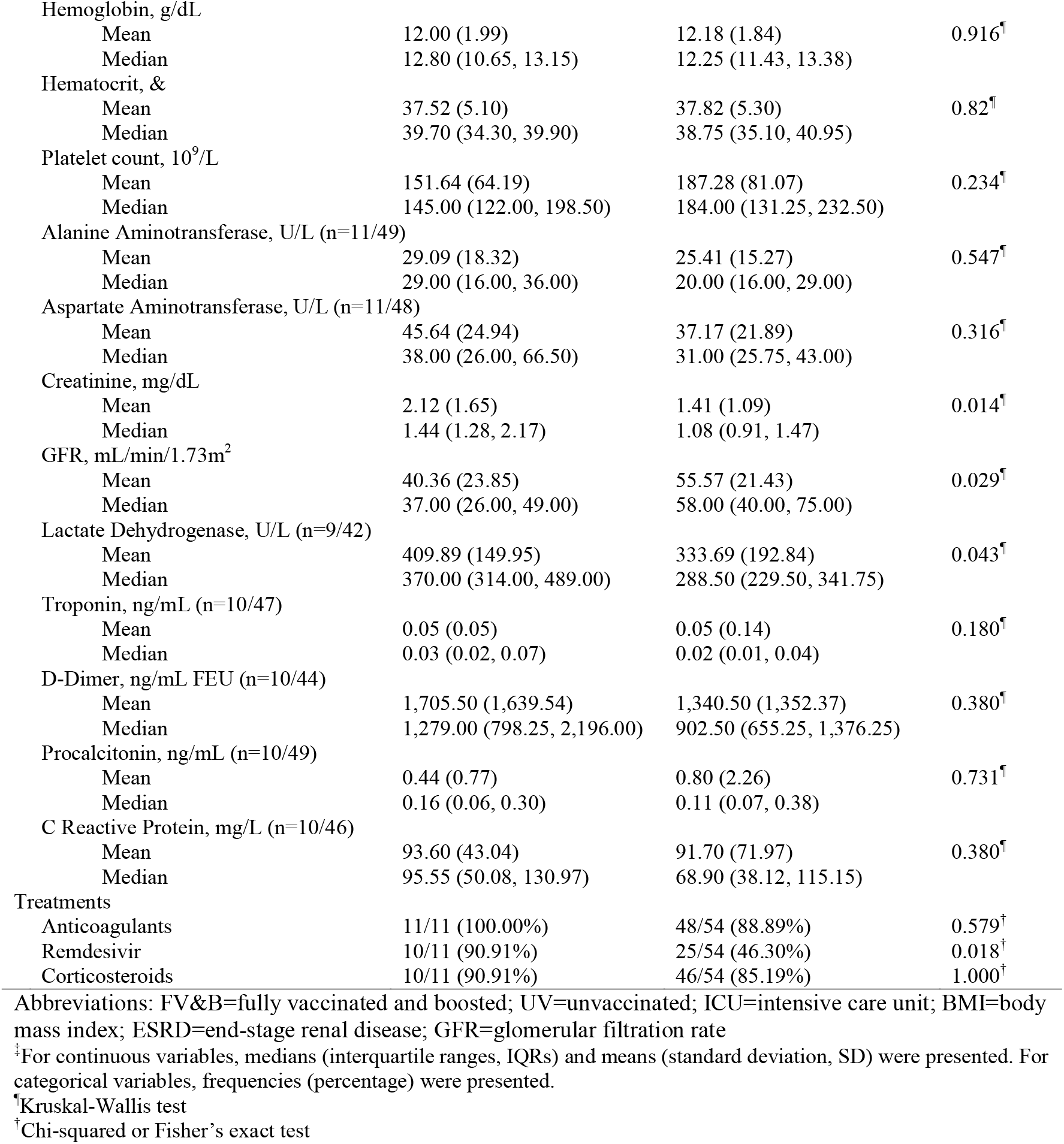
Past medical history, vital signs, laboratory values, and treatments among FV&B ICU patients and FV&B non-ICU patients

| Variables<sup>†</sup> | FV&B ICU | FV&B Non-ICU | *p* value |
| --- | --- | --- | --- |
| n | 11 | 54 |  |
| Past medical history |  |  |  |
| Elixhauser weighted score |  |  |  |
| Mean | 24.18 (13.67) | 14.35 (11.80) | 0.014<sup>¶</sup> |
| Median | 26.00 (19.50, 29.50) | 13.00 (5.00, 22.00) |  |
| Immunocompromised | 3/11 (27.27%) | 18/54 (33.33%) | 1.000<sup>‡</sup> |
| ED visit in prior 6 months | 3/11 (27.27%) | 15/54 (27.78%) | 1.000<sup>‡</sup> |
| COVID-19 symptoms |  |  |  |
| Respiratory and/or systemic symptoms | 9/11 (81.82%) | 50/54 (92.59%) | 0.266<sup>‡</sup> |
| Respiratory symptoms | 9/11 (81.82%) | 46/54 (85.19%) | 0.673<sup>‡</sup> |
| Cough | 7/11 (63.64%) | 35/54 (64.81%) | 1.000<sup>‡</sup> |
| Cough with sputum production | 1/11 (9.09%) | 2/54 (3.70%) | 0.432<sup>‡</sup> |
| Shortness of breath | 8/11 (72.73%) | 35/54 (64.81%) | 0.737<sup>‡</sup> |
| Sore throat | 0/11 (0.00%) | 4/54 (7.41%) | 1.000<sup>‡</sup> |
| Systemic | 5/11 (45.45%) | 38/54 (70.37%) | 1.000<sup>‡</sup> |
| Fever | 3/11 (27.27%) | 25/54 (46.30%) | 0.326<sup>‡</sup> |
| Fatigue | 5/11 (45.45%) | 30/54 (55.56%) | 0.742<sup>‡</sup> |
| Days from booster to symptom onset |  |  |  |
| Mean | 18.18 (21.53) | 40.70 (49.55) | 0.367<sup>¶</sup> |
| Median | 12.00 (7.50, 19.50) | 22.00 (5.00, 66.00) |  |
| ≥12 days | 6/11 (54.55%) | 32/54 (59.26%) | 1.000<sup>‡</sup> |
| Days from symptom onset to ED |  |  |  |
| Mean | 4.31 (3.20) | 4.64 (3.59) | 0.889<sup>¶</sup> |
| Median | 2.70 (1.80, 7.65) | 3.95 (1.70, 6.92) |  |
| Vaccine information |  |  |  |
| All Pfizer | 5/11 (45.45%) | 32/54 (59.26%) | 0.412<sup>‡</sup> |
| All Moderna | 4/11 (36.36%) | 17/54 (31.48%) |  |
| All Janssen | 2/11 (18.18%) | 3/54 (5.56%) |  |
| Mix | 0/11 (0.00%) | 2/54 (3.70%) |  |
| Vital signs on presentation to ED |  |  |  |
| Systolic blood pressure, mmHg |  |  |  |
| Mean | 143.18 (31.19) | 134.81 (18.25) | 0.211<sup>¶</sup> |
| Median | 149.00 (129.50, 165.50) | 134.00 (119.25, 148.00) |  |
| Diastolic blood pressure, mmHg |  |  |  |
| Mean | 76.00 (13.30) | 70.37 (11.48) | 0.192<sup>¶</sup> |
| Median | 72.00 (68.00, 87.00) | 69.00 (62.00, 77.75) |  |
| Pulse, beats per minute |  |  |  |
| Mean | 93.36 (17.41) | 92.26 (17.36) | 0.630<sup>¶</sup> |
| Median | 90.00 (82.00, 107.50) | 87.50 (80.00, 102.75) |  |
| Respiratory rate, breaths per minute |  |  |  |
| Mean | 24.55 (6.68) | 20.46 (4.19) | 0.023<sup>¶</sup> |
| Median | 24.00 (20.00, 27.50) | 20.00 (18.00, 21.50) |  |
| Temperature, °F |  |  |  |
| Mean | 98.62 (1.21) | 98.73 (1.41) | 0.847<sup>¶</sup> |
| Median | 98.70 (97.90, 99.25) | 98.60 (97.93, 99.35) |  |
| Blood oxygen saturation, % |  |  |  |
| Mean | 90.36 (8.45) | 95.98 (3.32) | 0.041<sup>¶</sup> |
| Median | 95.00 (83.50, 96.50) | 97.00 (95.00, 98.00) |  |
| Laboratory findings |  |  |  |
| White blood cell count, 10<sup>9</sup>/L |  |  |  |
| Mean | 21.16 (37.19) | 6.69 (3.27) | 0.073<sup>¶</sup> |
| Median | 7.40 (6.45, 13.20) | 6.30 (4.62, 8.17) |  |
| Hemoglobin, g/dL |  |  |  |
| Mean | 12.00 (1.99) | 12.18 (1.84) | 0.916<sup>¶</sup> |
| Median | 12.80 (10.65, 13.15) | 12.25 (11.43, 13.38) |  |
| Hematocrit, & |  |  |  |
| Mean | 37.52 (5.10) | 37.82 (5.30) | 0.82<sup>¶</sup> |
| Median | 39.70 (34.30, 39.90) | 38.75 (35.10, 40.95) |  |
| Platelet count, 10<sup>9</sup>/L |  |  |  |
| Mean | 151.64 (64.19) | 187.28 (81.07) | 0.234<sup>¶</sup> |
| Median | 145.00 (122.00, 198.50) | 184.00 (131.25, 232.50) |  |
| Alanine Aminotransferase, U/L (n=11/49) |  |  |  |
| Mean | 29.09 (18.32) | 25.41 (15.27) | 0.547<sup>¶</sup> |
| Median | 29.00 (16.00, 36.00) | 20.00 (16.00, 29.00) |  |
| Aspartate Aminotransferase, U/L (n=11/48) |  |  |  |
| Mean | 45.64 (24.94) | 37.17 (21.89) | 0.316<sup>¶</sup> |
| Median | 38.00 (26.00, 66.50) | 31.00 (25.75, 43.00) |  |
| Creatinine, mg/dL |  |  |  |
| Mean | 2.12 (1.65) | 1.41 (1.09) | 0.014<sup>¶</sup> |
| Median | 1.44 (1.28, 2.17) | 1.08 (0.91, 1.47) |  |
| GFR, mL/min/1.73m<sup>2</sup> |  |  |  |
| Mean | 40.36 (23.85) | 55.57 (21.43) | 0.029<sup>¶</sup> |
| Median | 37.00 (26.00, 49.00) | 58.00 (40.00, 75.00) |  |
| Lactate Dehydrogenase, U/L (n=9/42) |  |  |  |
| Mean | 409.89 (149.95) | 333.69 (192.84) | 0.043<sup>¶</sup> |
| Median | 370.00 (314.00, 489.00) | 288.50 (229.50, 341.75) |  |
| Troponin, ng/mL (n=10/47) |  |  |  |
| Mean | 0.05 (0.05) | 0.05 (0.14) | 0.180<sup>¶</sup> |
| Median | 0.03 (0.02, 0.07) | 0.02 (0.01, 0.04) |  |
| D-Dimer, ng/mL FEU (n=10/44) |  |  |  |
| Mean | 1,705.50 (1,639.54) | 1,340.50 (1,352.37) | 0.380<sup>¶</sup> |
| Median | 1,279.00 (798.25, 2,196.00) | 902.50 (655.25, 1,376.25) |  |
| Procalcitonin, ng/mL (n=10/49) |  |  |  |
| Mean | 0.44 (0.77) | 0.80 (2.26) | 0.731<sup>¶</sup> |
| Median | 0.16 (0.06, 0.30) | 0.11 (0.07, 0.38) |  |
| C Reactive Protein, mg/L (n=10/46) |  |  |  |
| Mean | 93.60 (43.04) | 91.70 (71.97) | 0.380<sup>¶</sup> |
| Median | 95.55 (50.08, 130.97) | 68.90 (38.12, 115.15) |  |
| Treatments |  |  |  |
| Anticoagulants | 11/11 (100.00%) | 48/54 (88.89%) | 0.579<sup>†</sup> |
| Remdesivir | 10/11 (90.91%) | 25/54 (46.30%) | 0.018<sup>†</sup> |
| Corticosteroids | 10/11 (90.91%) | 46/54 (85.19%) | 1.000<sup>†</sup> |

Abbreviations: FV&B=fully vaccinated and boosted; UV=unvaccinated; ICU=intensive care unit; BMI=body mass index; ESRD=end-stage renal disease; GFR=glomerular filtration rate
<sup>‡</sup>For continuous variables, medians (interquartile ranges, IQRs) and means (standard deviation, SD) were presented. For categorical variables, frequencies (percentage) were presented.
<sup>¶</sup>Kruskal-Wallis test
<sup>†</sup>Chi-squared or Fisher's exact test

Admission vital signs were similar between FV&B groups requiring ICU and non-ICU care except for slight differences in respiratory rate and oxygen saturation. Initial laboratory values were similar between groups except for renal function and lactate dehydrogenase levels. Patient’s requiring ICU-level care had a lower calculated glomerular filtration rate (GFR) (37.0 [26.0, 49.0] mL/min/1.73m^2^ vs 58.0 [40.0, 75.0] mL/min/1.73m^2^; p=0.03) and higher lactate dehydrogenase 370 (314, 489) U/L vs 288.5 (229.5, 341.8) U/L. The median Elixhauser Comorbidity Index was 26 (19.5, 29.5) for ICU patients, compared to 13 (5.0, 22.0) for non-ICU patients (p=0.01). 35 (53.8%) of all patients received Remdesivir; 10 (90.9%) in the ICU population vs 25 (46.3%) in the non-ICU population. 3/11 (27.3%) of FV&B patients in the ICU were immunocompromised vs 18/54 (33.3%) of non-ICU FV&B patients. **(Table 3)**

In the FV&B cohort that required ICU-level care (n=11), 7 (63.6%) were male with a median age of 71 (66.5, 76.5). 5 (45.5%) were vaccinated with three consecutive Pfizer immunizations, 4 (36.4%) received three doses of the Moderna vaccine, and 2 (18.2%) received two Janssen vaccines. Overall, among the 11 FV&B patients who required ICU level of care, 4 (36.4%) died, 3 (27.2%) were discharged to a Skilled Nursing Facility (SNF), 2 (18.2%) were discharged to a rehabilitation center, and 2 (18.2%) were discharged home. Median time interval from booster to onset of COVID-19 symptoms was 12 days (7.5, 19.5) with (5/11) 45% in less than 12 days. **(Figure 2)**

**Figure 2.**
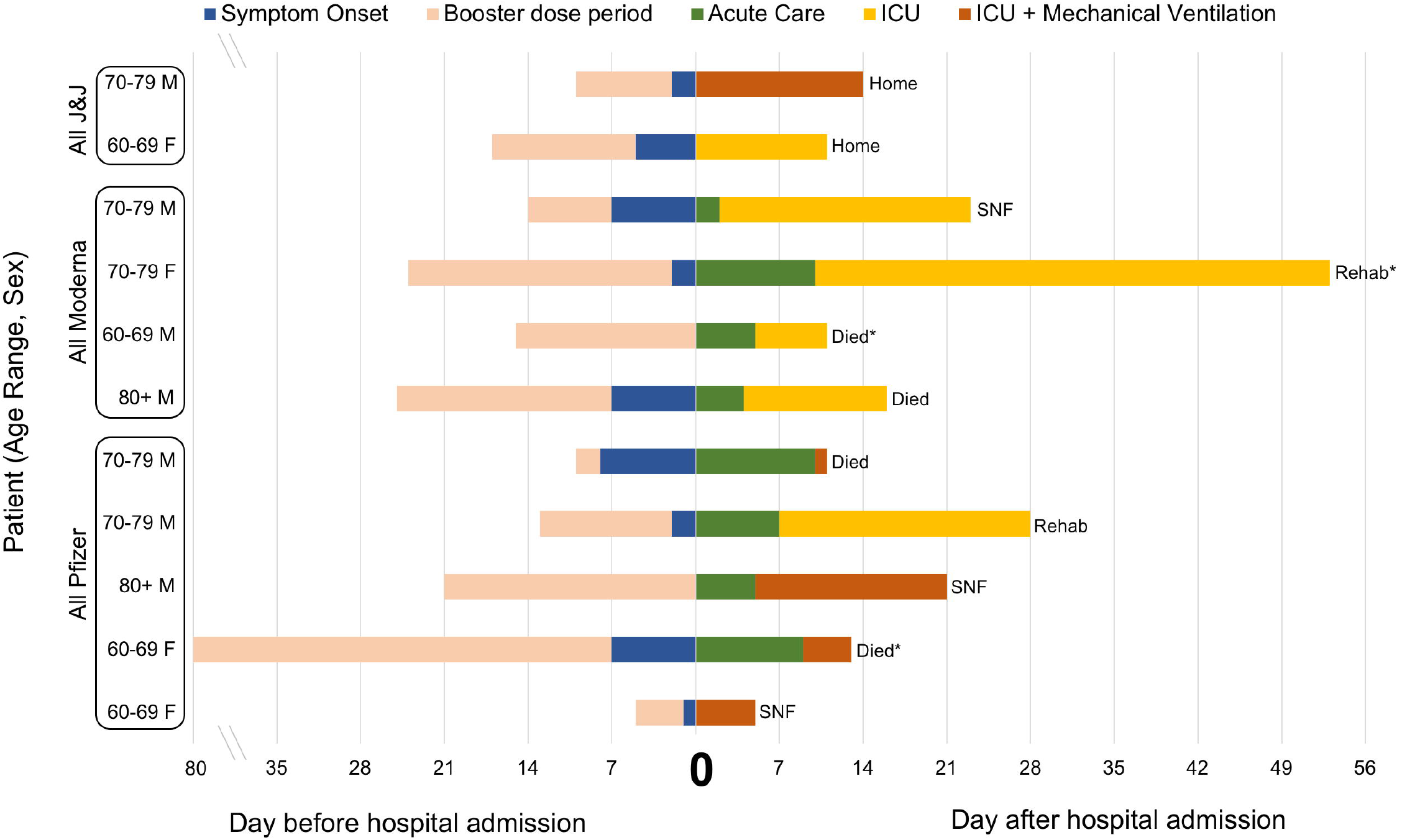
Characteristics and outcomes of 11 fully vaccinated and boosted patients with COVID-19 requiring intensive care admission. Demographic/clinical data includes age, sex, immunocompromised state, vaccine type, hospitalization units, timing of booster, and timing of symptom onset. Outcomes data includes disposition from hospital, mechanical ventilation, and mortality. ICU=Intensive care unit; SNF= Skilled nursing facility; Rehab= Rehabilitation *Immunocompromised

## LIMITATIONS

There were several limitations in this study. First, the sample size of the FV&B group was small. This was due to a smaller proportion of the population that was eligible for the booster dose during the study period. Further, given that our study only evaluated patients who sought emergency care and subsequently required hospitalization, with presumed strengthened immunity from the additional dose, there is likely a number of breakthrough SARS-CoV-2 infections that were not captured in our cohort. Given the staged approval of the booster doses starting with high-risk individuals, our sample population was biased towards individuals ≥ 65 years old as well as those with a pre-existing immunocompromised state. As with all retrospective studies, we were unable to control for confounders that may have affected hospital outcomes when comparing FV&B to UV individuals. Lastly, given our small sample size and the heterogeneity of vaccination sequences we were unable to determine if one vaccine, or sequence of vaccines, has benefit over another.

## DISCUSSION

Our study is one of the first investigations of the real-world effectiveness of booster dose therapy in combating severe COVID-19 in the United States. Despite older age (median 74 vs 58 years old), higher rate of pre-existing ESRD (18.5% vs 1.8%), higher proportion of immunocompromised state (32.3% vs 10.4%), and a higher risk of in-hospital death (Elixhauser 16 vs 8 (p <0.001)), in-hospital mortality trended lower in the FV&B vs UV group (7.7% vs 12.1%). While it is unsurprising that our FV&B cohort was older and had more immunocompromised individuals given the staged rollout of the booster vaccine by age and risk factors, it is notable that they experienced a lower rate of in-hospital mortality despite being higher risk for death at baseline. Given the Elixhauser comorbidity score of each group (16 for FV&B and 8 for UV) we would have anticipated approximately an 11% mortality rate in the FV&B cohort and a 5% mortality rate in the UV group at baseline.^20^ Although our observed difference in mortality did not reach statistical significance, given that we anticipated a higher mortality rate in this higher risk FV&B population, the trend towards a lower in-hospital mortality among that group is promising. Previous evidence highlights that severe outcomes amongst fully vaccinated individuals who are elderly with multiple comorbidities did not gain a significant mortality benefit once breakthrough infection requiring hospitalization had occurred.^21^ In this analysis, we saw a trend that even among elderly patients with significant comorbidities, a booster vaccine may provide additional protection against the progression toward severe disease despite breakthrough infection requiring hospitalization. Specifically, we observed a trend toward less need for mechanical ventilation and less need for vasopressor support in the FV&B population. While only marginally statistically significant given the small sample size, the mortality difference of 13.7% between FV&B and UV individuals over 65 years old was also eye-opening. While these findings are promising, additional analysis in larger trials is needed to account for the baseline level of illness in these two groups along with outcomes to determine if this trend is truly valid. However, this current analysis suggests an additional value of the booster that was not seen in previous literature on fully vaccinated patients and severe disease.^21,22^

The relationship between the timing of booster administration and the onset of infectious symptoms is notable in the FV&B ICU population. In general, COVID-19 was diagnosed in close proximity to receiving the dose. Specifically, 45.5% of the ICU group received the booster dose <12 days from onset of symptoms suggesting that FV&B patients who required ICU care may not have had enough time since their booster dose to confer full benefit of the additional vaccination. In fact, only one ICU patient had symptom onset 30 days after receiving a booster dose. The relevance of the timing of booster administration is supported by data from an Israeli study assessing the benefit of BNT162b2 booster. In that analysis, the authors found that 12 days post-booster immunization, there was a significant decrease in the rate of breakthrough infection when compared to the control, un-boosted, group.^23^ When comparing the FV&B ICU and non-ICU cohorts in our analysis, the median number of days from booster to symptom onset was 12 days in the ICU group compared to 22 days in the non-ICU group, further highlighting the delay in full potency of the booster in preventing severe disease.

It is unclear if an immunocompromised state is an independent predictor of severe COVID-19 disease in boosted patients. In our study sample, the proportion of individuals who were identified as immunocompromised was actually lower (27.3% vs 33.3%) in the patients who required ICU care. Previous evidence has shown that immunocompromised patients may have a tempered immune response to vaccination.^24,25^ However, additional doses may increase the likelihood of immune response.^26^ Interestingly, two ICU patients in our cohort had antibody testing (one anti-spike and one anti-nucleocapsid) at the time of presentation and neither had detectable antibody levels.

## CONCLUSIONS

In this study of hospitalized patients with COVID-19, we observed that the cohort of FV&B patients trended toward lower rates of mechanical ventilation, use of vasopressors, and in-hospital mortality compared to the cohort of UV patients, despite being higher risk for in hospital-mortality at baseline. This trend suggests that booster therapy may provide an additional protective benefit that was not seen in prior investigations evaluating breakthrough COVID-19 requiring hospitalization. Furthermore, when comparing FV&B patients who required ICU-level care against those who did not, ICU patients had on average less days since booster dose to symptom onset, suggesting that they may not have had enough time for the additional dose to have an effect. As COVID-19 continues to spread, more extensive studies are needed to further identify risk factors for severe outcomes among the FV&B population.

## Data Availability

The data that support the findings of this study are available via a data access agreement. Please contact the corresponding author (AB) for this request.

## AUTHOR CONTRIBUTIONS

AB, NM, and SJ designed the study, had full access to the data, and take responsibility for the integrity and accuracy of the data analysis. AB, NM, and SJ contributed to subject enrollment, data collection, data and statistical analysis. All authors contributed to the writing and editing of the manuscript. All authors contributed to data acquisition, analysis and interpretation, and all reviewed and approved the final version of the manuscript. The corresponding author attests that all listed authors meet authorship criteria and that no others meeting the criteria have been omitted.

## ETHICS COMMITTEE APPROVAL

This study was approved by the Beaumont Health Institutional Review Board.

